# Integrated analysis of bulk multi omic and single-cell sequencing data confirms the molecular origin of hemodynamic changes in Covid-19 infection explaining coagulopathy and higher geriatric mortality

**DOI:** 10.1101/2020.04.26.20081182

**Authors:** Shreya Johri, Deepali Jain, Ishaan Gupta

## Abstract

Besides severe respiratory distress, recent reports in Covid-19 patients have found a strong association between platelet counts and patient survival. Along with hemodynamic changes such as prolonged clotting time, high fibrin degradation products and D-dimers, increased levels of monocytes with disturbed morphology have also been identified. In this study, through an integrated analysis of bulk RNA-sequencing data from Covid-19 patients with data from single-cell sequencing studies on lung tissues, we found that most of the cell-types that contributed to the altered gene expression were of hematopoietic origin. We also found that differentially expressed genes in Covid-19 patients formed a significant pool of the expressing genes in phagocytic cells such as Monocytes and platelets. Interestingly, while we observed a general enrichment for Monocytes in Covid-19 patients, we found that the signal for FCGRA3+ Monocytes was depleted. Further, we found evidence that age-associated gene expression changes in Monocytes and platelets, associated with inflammation, mirror gene expression changes in Covid-19 patients suggesting that pro-inflammatory signalling during aging may worsen the infection in older patients. We identified more than 20 genes that change in the same direction between Covid-19 infection and aging cells that may act as potential therapeutic targets. Of particular interest were IL2RG, GNLY and GMZA expressed in platelets, which facilitates cytokine signalling in Monocytes through an interaction with platelets. To understand whether infection can directly manipulate the biology of Monocytes and platelets, we hypothesize that these non-ACE2 expressing cells may be infected by the virus through the phagocytic route. We observed that phagocytic cells such as Monocytes, T-cells, and platelets have a significantly higher expression of genes that are a part of the Covid-19 viral interactome. Hence these cell-types may have an active rather than a reactive role in viral pathogenesis to manifest clinical symptoms such as coagulopathy. Therefore, our results present molecular evidence for pursuing both anti-inflammatory and anticoagulation therapy for better patient management especially in older patients.

## Introduction

Coronavirus disease 2019 (Covid-19) caused by Severe acute respiratory syndrome (SARS)-coronavirus-2 (SARS-COV-2) is responsible for the ongoing pandemic that began in late December in Wuhan, China. Clinically, it is an acute respiratory illness which is resolved in more than 80% of cases. However about ∼ 20% of patients develop severe disease with a high fatality rate especially among aged patients1. It has been observed that non-survivors rapidly develop acute respiratory distress syndrome (ARDS) and multiple organ dysfunction syndrome (MODS) due to micro and macro circulatory thrombosis^1,2^. Disease is mostly transmitted by aerosols and patients have mild to severe pneumonia with bilateral chest infiltrates observed in chest imaging. Histopathologically, autopsy studies show bilateral diffuse alveolar damage. The interstitium shows inflammatory cells rich in lymphocytes, plasma cells, monocytes with giant cells. In addition to pulmonary pathology, Covid-19 afflicted patients have significant hemodynamic derangements at the severe end of the disease spectrum3. These include lymphopenia, thrombocytopenia, and monocytopenia with morphologically abnormal monocytes in the peripheral circulation. Coagulation work-up of these patients show deranged coagulation tests and pro-thrombotic profile of coagulation parameters in these patients. Non-survivor patients develop disseminated intravascular coagulation (DIC) in more than 70% of cases 4. These high venous thromboembolism rates were observed all across the body.

Virchow’s triad5 is the key to pathogenesis of thrombosis which includes 3 components: (1) Endothelial injury, (2) stasis or turbulent blood flow, and (3) hypercoagulability of the blood. While the latter two are related to circulatory disturbances and coagulation disorders, endothelial injury is caused by inflammation which invariably leads to platelet activation and thrombus formation. SARS-COV-2 causes thrombocytopenia either by directly affecting marrow cells or by activating the immune system to produce auto-antibodies against platelets^6^. During the process of lung injury, the virus increases platelet consumption by activation of platelets, platelet aggregation and wrapping of platelets. While megakaryocyte fragmentation in injured pulmonary bed causes reduced platelet production. A meta-analysis of 9 studies show an inverse correlation of platelet count with severity and greatly enhanced risk of mortality in Covid-19 patients7. Further, a longitudinal study found that enhanced platelet counts were significantly correlated with better prognosis8. Hence, it has been advised to monitor these patients by serial platelet count during their hospitalization. Abnormal coagulation is another complication of Covid-19 similar to other viral infections which is clinically manifested by intravascular thrombosis and DIC9. On the other hand, monocytes are infected by Covid-19 possibly through ACE2 receptors10. These infected monocytes cause inflammatory activation and produce a cytokine-storm^10,11^. This results in differentiation of monocytes into macrophages and they may show abnormal morphologic changes in peripheral circulation which are detected by flow cytometry examination. During the severe disease, these monocytes migrate to lungs and other affected organs causing monocytopenia^12^. Hence mounting evidence has confirmed the participation of platelets, endothelial cells and monocytes in pathogenesis and resultant clinical presentation in Covid-19 patients. However, the molecular players involved in these processes are unidentified.

In order to identify these molecular players, researchers have generated a few multi-omic datasets quantifying viral-host interactions. Two studies have elucidated the effect of Covid-19 infection on the bulk gene expression in lungs of deceased patients^13^ and peripheral blood monocytes (PBMCs)^14^ of active patients using RNA-sequencing and proteomics to identify unique transcriptional signatures in Covid-19 that correlate with clinical observations of host inflammatory responses such as cytokine-storms. Another study used the above bulk gene expression measurements to identify the gene expression changes in patient PBMCs to evaluate the effect of Hydroxychloroquine (HQ)^15^. Further, researchers have used affinity-purification mass spectrometry to quantify the protein-protein interactions between viral proteins and human host proteins. These studies found 553 human host proteins in a human kidney cell line (HEK293) that suggest the mechanism of how Covid-19 invokes host inflammatory response^14^ and identified potential drug targets for therapy^16^.

Most human tissues are composed of several functionally distinct cell-types defined by their unique gene expression signature. Recent international efforts such as the Human Cell Atlas^17^ have published detailed transcriptomic signatures of various individual cells across various tissues which can be used to dissect the cell-type specificity of the Covid-19 specific gene expression changes and viral-host interaction towards a better understanding of disease pathogenesis and progression. Leveraging the datasets generated in these studies, we searched for cell-type specific signatures in bulk RNA-sequencing of lung tissues from Covid-19 patients. Our investigation of disease-associated gene expression changes point towards age-associated priming of the platelet immune axis, explaining higher infectivity of Covid-19 in older patients. Further, we find evidence that non-ACE2 expressing cells, such as monocytes, dendritic cells and platelets, have a higher enrichment of genes that physically interact with the Covid-19 virus proteome. Interestingly most of these cells phagocytose infected cells or pathogens suggesting that the Covid-19 virus may be directly infecting these through phagocytosis to manipulate their biology towards an active participation in Covid-19 infection.

## METHODS

### Bulk gene expression deconvolution

De-convolution of gene expression from published bulk RNA-Seq dataset from lungs of healthy and Covid-19 patients was carried out using the a co-variance weighted gene expression single-cell transcriptomic deconvolution method published previously and available as the ‘MuSiC’ R package^18^

### Single-cell analysis using Seurat

Seurat^19^ package was used for single cell analysis. Data cleanup was done, and cells with very few genes (<0.05 quantile) and high mitochondrial gene expression (>0.95 quantile) were removed to exclude low quality and dying cells. Following cleanup, global scaling normalization was done such that the feature expression measurements for each cell were normalized by the total expression and then log transformed. As a prior to dimensionality reduction step using Principal Component Analysis (PCA), the data was scaled to a normal distribution. Dimensionality of the dataset was determined using the JackStraw procedure and the first 10 PCs were used for further analysis. The cells were subsequently clustered using graph-based clustering approaches, where each cell was embedded into a graph and edges between two cells were based on similar gene expression. Highly interconnected communities were identified as clusters.

### Published Datasets used

The following datasets were used for the current analysis in the specified form.

#### 1. Single cell RNA-Sequencing data

a. Pre-processed single cell dataset for lung, Oesophagus and Spleen^20^ were used with minimal modifications due to extremely useful formatting and excellent data annotation. This data was downloaded from: https://www.tissuestabilitycellatlas.org/
b. One pre-processed data set for 2.7k PBMCs was downloaded from a Seurat tutorial vignette https://www.dropbox.com/s/63gnlw45jf7cje8/pbmc3k_final.rds?dl=1 and used as it is.

#### 2. RNA Sequencing data from Covid-19 patient lung

The count matrices of Healthy and Covid-19 lung patients from Blanco-Melo et al ^13^ were downloaded from Gene expression Omnibus Dataset ID. **GSE147507**.

#### 3. RNA Sequencing data from platelets from Aging

The count matrices for the data were kindly provided by Dr. Cambell and Dr. Rondina from their paper exploring gene expression signatures of aging Human platelets ^21^.

#### 4. Viral-Host Protein interactome datasets

Images taken from published studies by manually parsing Figure 2 from Li et al^14^ and Figure 3 from Gordon et al^16^ available as Table 1 and Table 2 respectively.

**Figure 1:**
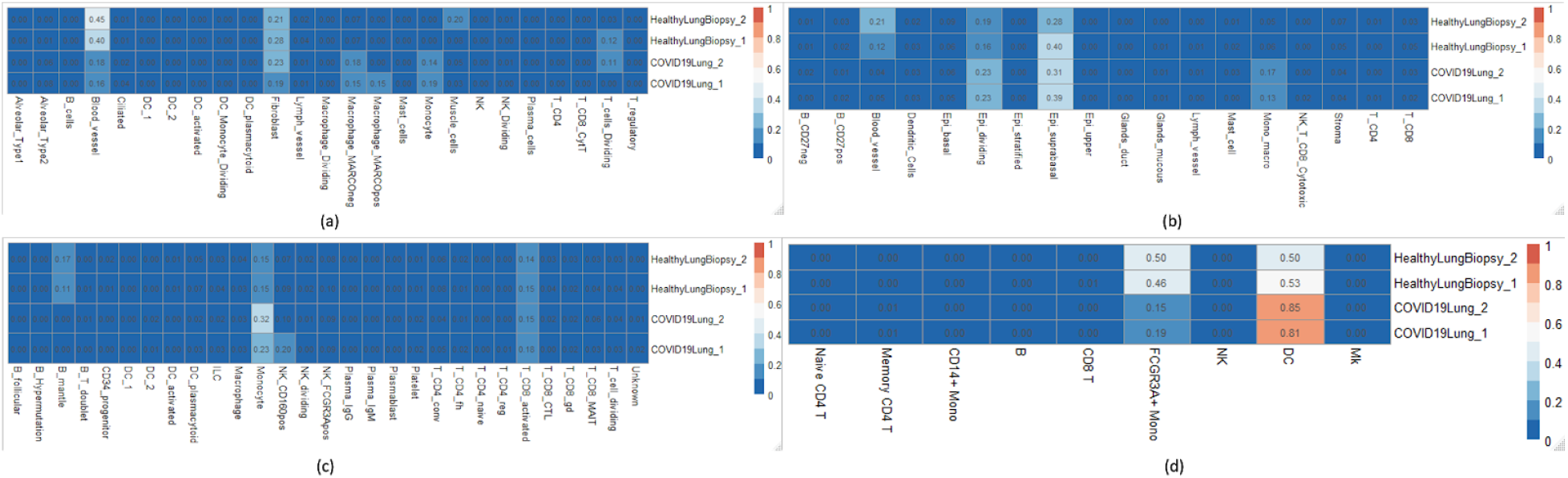
Cell-type specific signatures in Covid-19 patient lung bulk RNA-Seq. Deconvolution with (a) sc-RNA lung data shows enhanced gene expression for Monocytes and Macrophages and diminished gene expression for Blood vessels, (b) with sc-RNA oesophagus data shows enhanced gene expression for Monocytes and Epiblast cells and diminished gene expression for Blood vessels for Covid-19 patients, (c) with sc-RNA PBMC data shows enhanced gene expression for Dendritic cells and strongly diminished gene expression for FCGR3A+ Monocytes, and (d) with sc-RNA Spleen shows diminished gene expression for B-mantle cells in Covid-19 patients.

**Figure 2:**
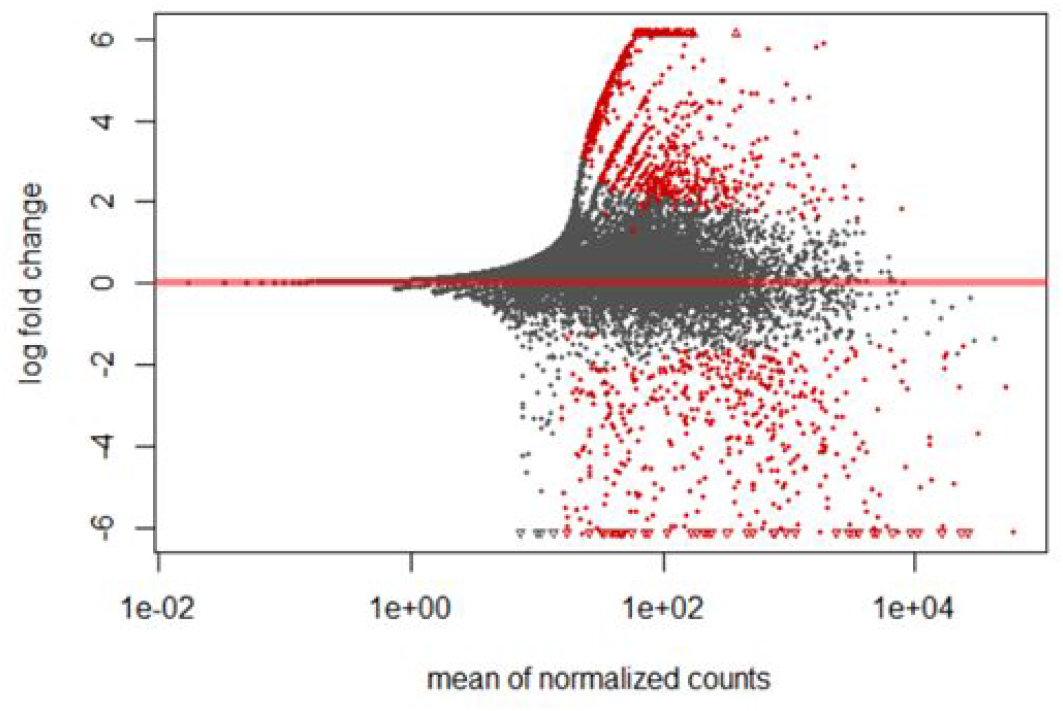
MA Plot for differentially expressed genes. Shows the log2 fold changes attributable to a given variable over the mean of normalized counts for all the samples. Red colored points have FDR <0.05

**Figure 3:**
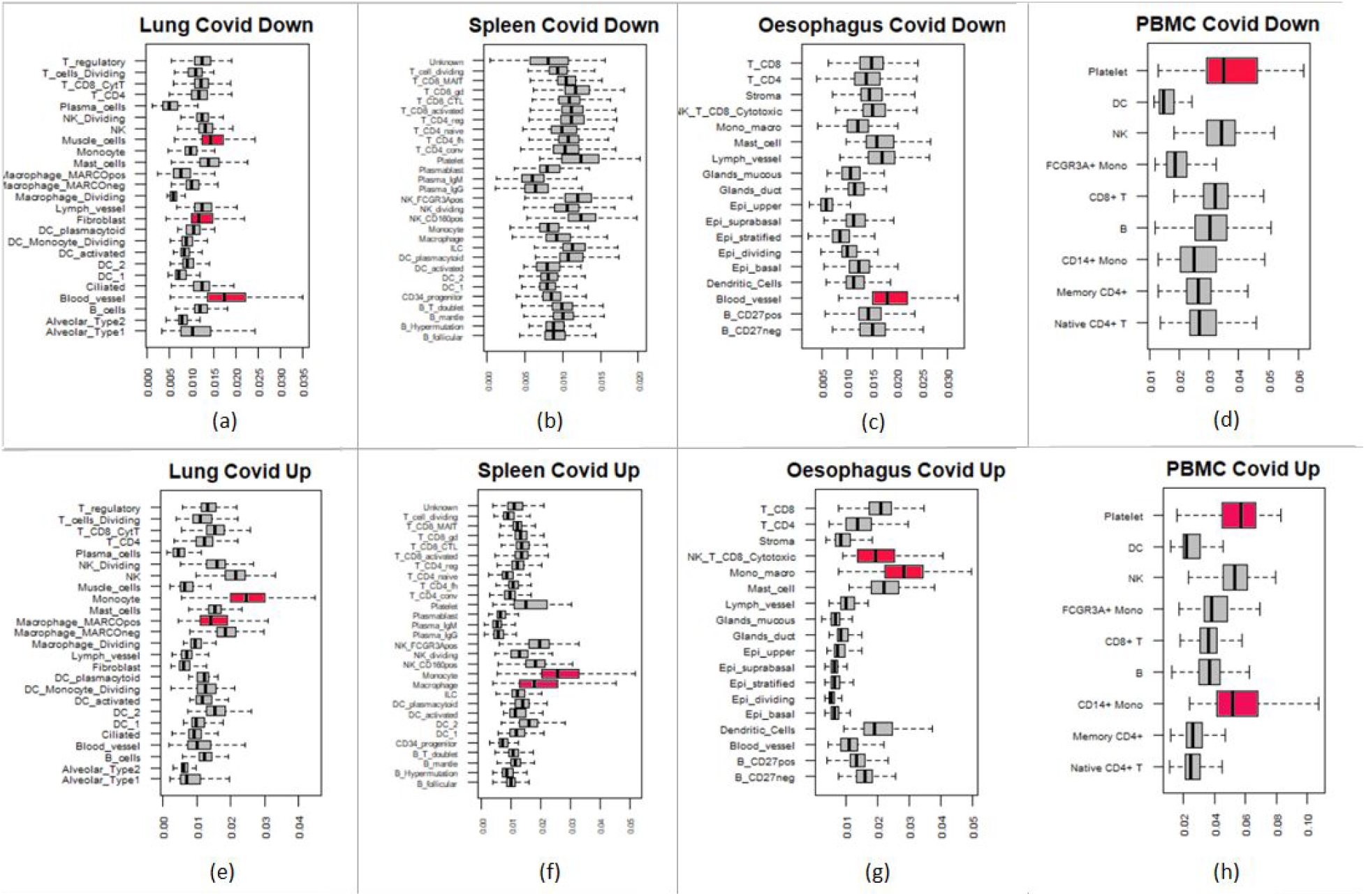
Cell-type specific enrichment of differentially expressed genes in Covid-19 Patient Lung tissue. (a) Genes downregulated in Covid-19 enriched in Blood vessels, Muscle cells and Fibroblasts of Lungs, (b) no specific type in Spleen, (c) Blood vessels of Oesophagus, and (d) Platelets in PBMC. Genes upregulated in Covid-19 enriched in (e) Monocytes and Macrophages of Lungs, (f) Macrophages and Monocytes in Spleen, (g) Monocytes and Natural Killer CD8+ Cytotoxic cells in Oesophagus, and (h) Platelets and CD14+ Monocytes in PBMC.

## RESULTS

### Deconvolution of cell-type specific signals from bulk Covid-19 Patient lung tissue

In order to dissect the role of the above various cell-types found in the lungs and upper respiratory tract cells in Covid-19, we reasoned that the gene expression signature of some of these cells would be enriched in the bulk-RNA sequencing data from infected patients samples in comparison with healthy controls. We employed weighted bulk RNA-Sequencing gene expression deconvolution method MuSiC^18^ on data from lung tissue of two healthy and two Covid-19 patients^13^ (Figure 1). By deconvoluting bulk tissue expression using single-cell transcriptomics data from lung tissues, we found that the gene expression signature for Macrophages and Monocytes was enhanced while that of Blood vessels was diminished in the Covid-19 patients. This correlates with clinical observations and suggests a possible direct role of the virus in immune activation and destruction of blood vessels. As a sanity check, we also deconvolved bulk RNA-sequencing data using single-cell transcriptomic data from Oesophagus, due to its proximity to and compositional similarity with the upper respiratory tract, and found enhanced signature for Monocytes and diminished signature for Blood vessels further supporting our observation from lungs. We also found an enhanced gene expression signature for dividing epiblast cells in Covid-19 patients suggesting bodies’ response to replenish the epithelial cells destroyed upon infection. To further confirm the involvement of cells of the myeloid origin, we next used single-cell transcriptomics data from PBMCs for deconvolution. Interestingly here we found an enhanced gene-expression signature for Dendritic cells while a strongly diminished gene-expression signature for FCGRA3+ Monocytes in Covid-19 patients. FCGRA3+ Monocytes play a role in phagocytosis and interferon gamma response and a diminished signature indicates loss of these capabilities within Covid-19 patient lungs.

### Cell-type specific enrichment of differentially expressed genes in Covid-19 Patient lung tissue

To further gain evidence into the role of of specific cell-types and their sub-types towards Covid-19 pathology we performed differential expression analysis (Figure 2, 3) on bulk RNA-seq data from lung tissue of two healthy and two Covid-19 patients using DESeq2^22^ at an FDR of 5% to identify 419 down-regulated genes, enriched for circulatory and vascular development, and 395 upregulated genes, enriched for myeloid cell activation involved in immune response. Next, we calculated the percentage of total gene expression for the identified upregulated and downregulated genes in Covid-19 patients across constituent cell types in each of the 4 tissues - lung and Oesophagus representing the Lower and Upper Respiratory tract; while PBMCs and Spleen representing the cells of the hematopoietic origin and the Immune system. Echoing the results from the previous section, in lungs we found that the upregulated genes in Covid-19 patients were also enriched in Monocytes and Macrophages (p< 2e-16, in a one-way ANOVA) while the downregulated genes were enriched in Blood vessels, Muscle cells and Fibroblast cells (p< 2e-16, in a one-way ANOVA). This suggests that upon infection in lungs, the myeloid lineage of cells proliferate or accumulate from the peripheral circulatory system while the cells of epithelial origin forming structural framework disintegrate. However, in case of the Oesophagus, a proxy for the upper respiratory tract, besides Monocytes and Natural killer CD8+ T cells, blood vessels (p< 2e-16, in a one-way ANOVA) showing similar trends as in lungs. The upper epithelial cells (expressing Keratinocyte markers such KRT4 and ECM1) were particularly enriched (p< 2e-16, in a one-way ANOVA) for genes upregulated in Covid-19 patients. Interestingly, researchers found cells of epithelial origin to be more susceptible to environmental factors, resulting in higher mitochondrial gene expression indicative of cell death^20^. Therefore, an enhanced signature for epithelial cells could be indicative of their cell death in both the upper and lower respiratory tract due to the infection. In the case of PBMCs, we observe the CD14+ Monocytes and platelets (p< 2e-16, in a one-way ANOVA) enriched for genes upregulated in Covid-19 patients, while platelets (p< 2e-16, in a one-way ANOVA) are also enriched in genes downregulated in Covid-19 patients, indicating cell death as shown recently^23^. In the Spleen, Monocytes are enriched for genes upregulated in Covid-19 patients while no clear cell-type specific enrichment is seen among genes downregulated in Covid-19 patients. Results for the Tukey test for the same can be found in Supplementary Table 1.

These analyses lend further support to the role of the different Monocytes, T-lymphocytes and Blood vessels in Covid-19 pathogenesis. On deeper investigation of differentially regulated genes in Covid-19 patients we found the gene ADAMTS1, which had a 3 fold reduction in gene expression for Covid-19 patients with less than FDR 6%. ADAMTS1 gene plays a role in angiogenesis, coagulation and inflammation^24^ indicating that this could be a consequence of inflammation rather than a cause of it. Another explanation could be that due to the relatively smaller size and abundance of the platelets to contribute a significant amount of RNA molecules for a successful deconvolution of their gene expression signature from bulk tissue.

### Age-associated predisposition for Covid-19 infection through hemodynamic priming

Upon identification of cell-types playing a role in pathogenesis, we wanted to understand if age-associated gene expression changes in these very cell-types could predispose older patients to the virus by adopting gene expression signature observed in the Covid-19 patient lung samples in the previous section. In a study comparing Older versus Younger CD14+ Monocytes and CD4+ T-cells in more than 400 individuals using microarray expression data^25^, we found 19 genes that correlated with gene expression changes in Covid-19 patients. 8 genes (GBP5, CR1, ARRB2, CXCL16, MXD1, IL1R2, ACSL1, WAS) upregulated in Covid-19 patients had age-associated gene expression changes associated with gene expression changes in CD14+ Monocytes. While 13 genes (SMAP2, FCGR2A, SPI1, AQP9, CXCL16, C5AR1, FPR1, LILRB2, LILRA5, MXD1, FGL2, LYN, CYBB) upregulated in Covid-19 patients had age-associated associated gene expression changes in CD4+ T-cells. In another study following age-dependent gene expression response in blood upon infection with an influenza virus *ex-vivo*^26^, revealed 13 genes (CSF3R, S100A9, S100A8, FCGR2A, CR1, CLEC7A, ARHGDIB, CEACAM6, LILRB3, LILRA5, LILRA1, NCF4, TLR1, LY96) that were associated with age-dependent response and were upregulated in Covid-19 patients. Three genes CR1, Complement system receptor 1; FCGR2A, family of immunoglobulin Fc receptor; and LILRA5, regulating proinflammatory cytokine secretion, all found on the surface of Monocytes, were common between the two studies and could be potential drug targets. In another study comparing platelets from Old and Young humans^21^, we found that both Older patients and lungs of Covid-19 patients had the upregulation of IL2RG and GNLY, both proteins highly expressed on Natural Killer T-cells and play a role in cytokine signalling. Hence, the immune response by older platelets suggests enhanced cytokine signalling similar to Covid-19 infection based on similarity in gene expression. This in turn explains the clinical observations of coagulopathy in severe cases of Covid-19, and explains the role of platelets. In fact the platelet study also demonstrated that Granzyme A^21^, upregulated with older platelets, induces inflammation in Monocytes and cytokine secretion. Our analysis suggests that IL2RG and GNLY could mediate the interaction between platelets and Monocytes through Granzyme A which could act as potential therapeutic intervention that may alleviate symptoms including coagulopathy.

### Non-ACE2 expressing cells highly express proteins found in the viral proteome

Viruses hijack the human host cell machinery by interacting with its proteins. But viral entry into the host cell is a prerequisite for this to happen. In the case of Covid-19, viral entry was shown to be mediated by the ACE2 receptor and the TMPRSS2 protease^27^. However, PBMCs such as Macrophages, Monocytes, Dendritic cells, platelets and T-cells do not express either ACE2 or TMPRSS2 at a high level. Yet studies with the closely related SARS-CoV virus found that the virus can replicate in PBMC of patients^10,28^ although in pure Monocyte/Macrophage cultures different strains of the SARS-Cov virus replicate with a highly variable titre^10,28^. This non-ACE2 mediated entry into the cells can be explained due to phagocytosis as Monocytes/Macrophages, Dendritic cells, Neutrophils and even platelets ^29^ may give the Covid-19 virus entry by phagocytosing viral infected cells or directly phagocytosing the virus itself. To test this possibility, we hypothesized that cell-types which contribute the most to Covid-19 infection must be doing so by highly expressing genes that physically interact with viral proteins upon its entry.

Two recent studies capturing viral-host interactome tagged viral proteins as baits followed by affinity-purification and mass-spectrometry to identify a set of 332 (set 1: Gordon et al.^16^) and another set of 224 high confidence human proteins (set 2: Li et al.^14^) in HEK293 cell-lines which interact and co-precipitate with proteins of the Covid-19 (Supplementary Table 2). To test our hypothesis, we estimated the percentage expression of 553 host proteins (combined set of protein from both studies, Figure 4) that interact with viral proteins (a.k.a the viral interatome) across various cell-types in the different human tissues that may come in contact with the virus. As done in previous sections, we chose single-cell transcriptomics data from lungs, Oesophagus, PBMCs and Spleen (Figure 5). In lungs, high expression of the viral interactome was observed in NK cells and Blood vessels (p< 2e-16, in a one-way ANOVA); in the Oesophagus, high expression of the viral interactome was observed in Stromal cells, Monocytes and Blood vessels (p< 2e-16, in a one-way ANOVA); and in PBMCs, high expression of the viral interactome was observed in platelets/Megakaryocytes (p< 2e-16, in a one-way ANOVA) while in Spleen high expression of the viral interactome was observed in platelets (p< 2e-16, in a one-way ANOVA). Results for the Tukey test for the same can be found in Supplementary Table 3. These results of cell-type specific viral interactome gene expression enrichment confirm our hypothesis that viral proteins may be directly interacting with highly abundant proteins within Monocytes, platelets and Vasculature that induces cytokine signalling leading to the observed clinical manifestations observed in Covid-19. Thus, instead of alleviating symptoms the phagocytic activity of these cell-types may be further exacerbating the clinical symptoms. This has been observed in a number of other successful pathogens such as Gonorrhea and HIV.

**Figure 4:**
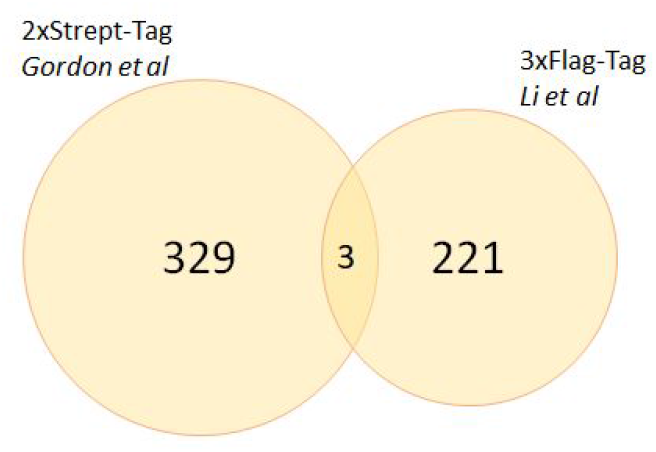
AP-MS captured Human proteins interacting with SARS-CoV-2 virus

**Figure 5:**
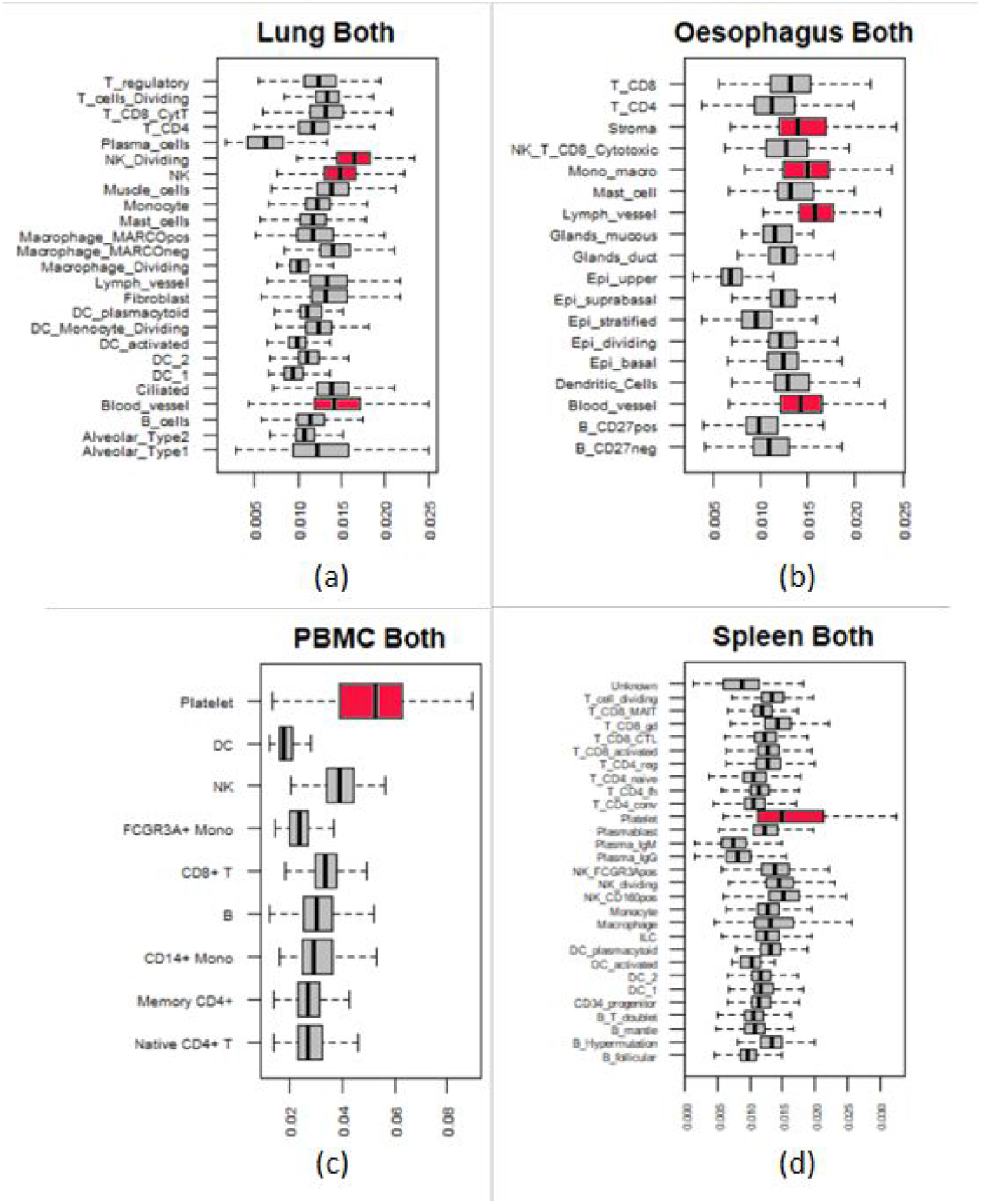
Expression of interactome proteins in different tissues. (a) In Lungs, the genes are highly expressed in NK cells, and Blood vessels, (b) in Oesophagus, they are highly expressed in Stroma cells, Monocytes, lymph and Blood vessels, and in PBMC (c) and Spleen (d), they are highly expressed in Platelets.

## DISCUSSION

Through computational methods of deconvolution and differential expression of the viral interactome proteins, we identified Monocytes, Natural killer T-cells, platelets and Blood vessels as the cell types that may be attacked by the SARS-CoV-2 viral proteins. We found that most of these cell types did not express the ACE2 receptor, hence the non-ACE2 mediated entry into the cells through the phagocytic pathway was suggested for a subset of these cells that phagocytose. Besides a role in inflammation, all these identified cell types also co-operate to play a role in coagulation explaining rampant observation of coagulopathy in severe Covid-19 patients. Hence, anticoagulants are being used during treatment. In a recent study, with 449 Covid-infected and 104 non Covid-infected patients, the role of Heparin, an anticoagulant, was studied. A lower mortality was observed in Heparin users as compared to nonusers when D-dimer concentration exceeded 3.0 μg/mL (32.8% vs. 52.4%, P=0.017). However, no difference was found in mortality rates between heparin users than nonusers in the non-COVID group when being stratified by D-dimer. Notably, the 28-day mortality of heparin users was lower than nonusers in patients^29^.

There are several limitations in the current study. First, the bulk-RNA seq lung data contained only two Covid positive patients and two Healthy controls. More robust results can be obtained if a larger dataset on Covid positive patients is available. Second, methods of deconvolution were used to extrapolate the bulk-RNA seq data to single cell level expression. We strongly suggest that detailed results can be obtained if single cell RNA-seq samples are available for patients infected with SARS-CoV-2.

We conclude our study with a few open questions. First, our study found evidence of an alternative mode of spread of the virus, through phagocytosis. Traditionally, phagocytosis is considered to be the step towards decimation of the infection, hence a deeper insight of the mechanism of spread through phagocytosis is yet to be found. Second, it has been suggested in some mice studies that older Hematopoietic Stem Cells (HSCs) are primed to make more platelets^30^. Other studies have demonstrated that the lungs are a site of platelet production^31^, which may also explain why lungs may be attacked vigorously by the virus by the route of ACE2 expressing epithelial cells and by even infecting the tissue resident platelet and Monocyte populations.

## Data Availability

The following datasets were used for the current analysis in the specified form.
Single cell RNA-Sequencing data:
Pre-processed single cell dataset for Lung, Oesophagus and Spleen20 were used with minimal modifications due to extremely useful formatting and excellent data annotation. This data was downloaded from:
https://www.tissuestabilitycellatlas.org/
One pre-processed data set for 2.7k PBMCs was downloaded from a Seurat tutorial vignette
https://www.dropbox.com/s/63gnlw45jf7cje8/pbmc3k_final.rds?dl=1 and used as it is.
RNA Sequencing data from Covid-19 patient Lung:
The count matrices of Healthy and Covid-19 Lung patients from Blanco-Melo et al were downloaded from Gene expression Omnibus Dataset ID. GSE147507.
RNA Sequencing data from Platelets from Aging:
The count matrices for the data were kindly provided by Dr. Cambell and Dr. Rondina from their paper exploring gene expression signatures of aging Human Platelets.
Viral-Host Protein interactome datasets:
Images taken from published studies by manually parsing Figure 2 from Li et al and Figure 3 from Gordon et al available as Table 1 and Table 2 respectively.

https://www.tissuestabilitycellatlas.org/

https://www.dropbox.com/s/63gnlw45jf7cje8/pbmc3k_final.rds?dl=1

## REFERENCES

1. Weiss, P. & Murdoch, D. R. Clinical course and mortality risk of severe COVID-19. The Lancet vol. 395 1014–1015 (2020).

2. Cannegieter, S. C. & Klok, F. A. COVID-19 associated coagulopathy and thromboembolic disease: Commentary on an interim expert guidance. Research and Practice in Thrombosis and Haemostasis (2020) doi: 10.1002/rth2.12350.

3. Yaqian, M., Lin, W., Wen, J. & Chen, G. Clinical and pathological characteristics of 2019 novel coronavirus disease (COVID-19): a systematic review. doi: 10.1101/2020.02.20.20025601.

4. Arachchillage, D. R. & Laffan, M. Abnormal coagulation parameters are associated with poor prognosis in patients with novel coronavirus pneumonia. J. Thromb. Haemost. (2020) doi: 10.1111/jth.14820.

5. Breddin, H. Thrombosis and Virchow’s Triad: What Is Established? Seminars in Thrombosis and Hemostasis vol. 15 237–239 (1989).

6. Xu, P., Zhou, Q. & Xu, J. Mechanism of thrombocytopenia in COVID-19 patients. Annals of Hematology (2020) doi: 10.1007/s00277-020-04019-0.

7. Thrombocytopenia is associated with severe coronavirus disease 2019 (COVID-19) infections: A meta-analysis. Clin. Chim. Acta 506, 145–148 (2020).

8. Liu, Y. et al. Association between platelet parameters and mortality in coronavirus disease 2019: Retrospective cohort study. platelets 1–7 (2020).

9. Zhang, W. et al. The use of anti-inflammatory drugs in the treatment of people with severe coronavirus disease 2019 (COVID-19): The Perspectives of clinical immunologists from China. Clin. Immunol. 214, 108393 (2020).

10. Yilla, M. et al. SARS-coronavirus replication in human peripheral monocytes/macrophages. Virus Res. 107, 93–101 (2005).

11. Mehta, P. et al. COVID-19: consider cytokine storm syndromes and immunosuppression. Lancet 395, 1033–1034 (2020).

12. Zhang, D. et al. COVID-19 infection induces readily detectable morphological and inflammation-related phenotypic changes in peripheral blood monocytes, the severity of which correlate with patient outcome. doi: 10.1101/2020.03.24.20042655.

13. Blanco-Melo, D. et al. SARS-CoV-2 launches a unique transcriptional signature from in vitro, ex vivo, and in vivo systems. doi: 10.1101/2020.03.24.004655.

14. Li, J. et al. Virus-host interactome and proteomic survey of PMBCs from COVID-19 patients reveal potential virulence factors influencing SARS-CoV-2 pathogenesis. doi: 10.1101/2020.03.31.019216.

15. Corley, M. J., Sugai, C., Schotsaert, M., Schwartz, R. E. & Ndhlovu, L. C. Comparative in vitro transcriptomic analyses of COVID-19 candidate therapy hydroxychloroquine suggest limited immunomodulatory evidence of SARS-CoV-2 host response genes. doi: 10.1101/2020.04.13.039263.

16. Mahen, R. A SARS-CoV-2-Human Protein-Protein Interaction Map Reveals Drug Targets and Potential Drug-Repurposing. doi: 10.1242/prelights.18355.

17. Regev, A. et al. The Human Cell Atlas. Elife 6, (2017).

18. Wang, X., Park, J., Susztak, K., Zhang, N. R. & Li, M. Bulk tissue cell type deconvolution with multi-subject single-cell expression reference. Nat. Commun. 10, 1–9 (2019).

19. Butler, A., Hoffman, P., Smibert, P., Papalexi, E. & Satija, R. Integrating single-cell transcriptomic data across different conditions, technologies, and species. Nat. Biotechnol. 36, 411–420 (2018).

20. Madissoon, E. et al. scRNA-seq assessment of the human lung, spleen, and esophagus tissue stability after cold preservation. Genome Biol. 21, 1 (2019).

21. Campbell, R. A. et al. Granzyme A in Human platelets Regulates the Synthesis of Proinflammatory Cytokines by Monocytes in Aging. J. Immunol. 200, 295–304 (2018).

22. Love, M. I., Huber, W. & Anders, S. Moderated estimation of fold change and dispersion for RNA-seq data with DESeq2. Genome Biology vol. 15 (2014).

23. Wang, X. et al. SARS-CoV-2 infects T lymphocytes through its spike protein-mediated membrane fusion. Cell. Mol. Immunol. (2020) doi: 10.1038/s41423-020-0424-9.

24. Kelwick, R., Desanlis, I., Wheeler, G. N. & Edwards, D. R. The ADAMTS (A Disintegrin and Metalloproteinase with Thrombospondin motifs) family. Genome Biology vol. 16 (2015).

25. Reynolds, L. M. et al. Transcriptomic profiles of aging in purified human immune cells. BMC Genomics 16, 333 (2015).

26. Piasecka, B. et al. Distinctive roles of age, sex, and genetics in shaping transcriptional variation of human immune responses to microbial challenges. Proc. Natl. Acad. Sci. U. S. A. 115, E488–E497 (2018).

27. Hoffmann, M. et al. SARS-CoV-2 Cell Entry Depends on ACE2 and TMPRSS2 and Is Blocked by a Clinically Proven Protease Inhibitor. 181, 271–280.e8 (2020).

28. Li L, E. al. SARS-coronavirus replicates in mononuclear cells of peripheral blood (PBMCs) from SARS patients. - PubMed - NCBI. https://www.ncbi.nlm.nih.gov/pubmed/14522061.

29. Matowicka-Karna, J., Kamocki, Z. & Kemona, H. Assessment of platelet activation and phagocytic activity in gastric cancer patients. World J. Gastrointest. Pathophysiol. 4, 12–17 (2013).

27. Li, Lanjuan, et al. “SARS-coronavirus replicates in mononuclear cells of peripheral blood (PBMCs) from SARS patients.” Journal of Clinical Virology 28.3 (2003): 239–244.

29. Yin, Shiyu, et al. “Difference of coagulation features between severe pneumonia induced by SARS-CoV2 and non-SARS-CoV2.” Journal of Thrombosis and Thrombolysis (2020): 1.

30. Frisch, Benjamin J., et al. “Aged marrow macrophages expand platelet-biased hematopoietic stem cells via interleukin-1B.” JCI insight 4.10 (2019).

31. Lefrançais, Emma, et al. “The lung is a site of platelet biogenesis and a reservoir for haematopoietic progenitors.” Nature 544.7648 (2017): 105–109.

